# Extracorporeal Blood Purification in moderate and severe COVID-19 patients: a prospective cohort study

**DOI:** 10.1101/2020.10.10.20210096

**Authors:** Rodney Alexander Rosalia, Petar Ugurov, Dashurie Neziri, Simona Despotovska, Emilja Kostoska, Lidija Veljanovska-Kiridjievska, Dimche Kuzmanov, Aleksandar Trifunovski, Gianluca Villa, Dijana Popevski, Zan Mitrev

## Abstract

**Introduction:** COVID-19 is characterised by hyperinflammation and coagulopathy. Severe cases often develop respiratory distress, requiring mechanical ventilation and critical cases progressing to ARDS. Control of hyperinflammation has been proposed as a possible therapeutic avenue for COVID-19; extracorporeal blood purification (EBP) modalities offer an attractive mean to ameliorate maladaptive inflammation.

With this work, we describe the longitudinal variation of parameters of systemic inflammation in critically ill COVID-19 patients treated with blood purification using AN69ST (oXiris^®^) hemodiafilter.

**Methods:** We performed a time-series analysis of 44 consecutive COVID-19 cases treated with the AN69ST (oXiris^®^) cytokine adsorbing hemodiafilter; we visualise longitudinal results of biochemical, inflammatory, blood gas- and vital sign parameters.

**Results:** Blood purification was indicated for suspected hyperinflammation or hypercoagulation, (= CRP ≥ 100 mg/L and/or IL-6 ≥ 40 pg/mL and/or Ferritin ≥ 500 ng/mL and/or Lactate Dehydrogenase > 365 U/L or D-dimers > 2000 ng/mL). All patients were treated with ≥ 1 cycle extracorporeal continuous venovenous hemofiltration (CVVHF) with cytokine adsorbing hemodiafilter (CAH); of these, 30 severe patients received CVVHF-CAH within 4 – 12 hours of hospitalisation. Another 14 patients admitted with mild-to-moderate symptoms progressed to severe disease and placed on EBP during the course of hospitalisation. The treatment was associated with a reduction of Ferritin, C-reactive protein, Fibrinogen, several inflammatory markers and a resolution of numerous cytopenias. The observed mortality across the cohort was 36.3% across the cohort.

**Conclusion:** Extracorporeal blood purification with cytokine adsorbing hemofilter was associated with a decrease in the acute phase proteins CRP, Ferritin, and resolution of numerous cytopenias. Repetitive hemofiltration has been associated with lower levels of IL-6 in COVID-19 patients.

## Introduction

The Coronavirus disease 2019 (COVID-19) pandemic dominated our lives since the outbreak of the Severe Acute Respiratory Syndrome Coronavirus 2 (SARS-CoV-2) started in Wuhan of Hubei Province, China [1]. Its disruptive nature was confirmed with the WHO’s classification as a global epidemic on February 28, 2020, [2].

Severe COVID-19 disease is characterised by uncontrolled inflammation [3-5]. COVID-19 patients typically present with bilateral multifocal peripheral lung changes of ground-glass opacity and, or consolidation, on chest radiography [6] low peripheral oxygen saturation (SpO_2_) and an increased respiratory rate strongly associated with pneumonia. Immunopathology [7, 8] is considered one of the main drivers of disease progression leading to acute respiratory distress syndrome (ARDS) [9], multiorgan failure [10, 11, 5], secondary infections [12] and coagulopathy [13, 14].

The use of blood purification devices has been advocated as a treatment option to mitigate atypical inflammation as a means to halt disease progression.

We here describe a case series of 44 COVID-19 patients treated with CVVHF-CAH using the AN69ST (oXiris®) filter and present longitudinal analysis of clinical and laboratory parameters associated with disease severity. EBP was supplemented with systemic heparinisation and non-invasive respiratory [15] support aimed at preventing irreversible lung pathology [16] and the necessity of invasive mechanical ventilation. Mechanical respiratory support is linked to high mortality rates, especially in older patients or those with severe comorbidities [17].

This prospective cohort study evaluated variation in several biomarkers, clinical parameters and outcome associated with EBP to control hyper inflammation. Institutional review board approval was obtained from the local ethical committee of the Zan Mitrev Clinic in accordance with the declaration of Helsinki and ICH-Good Clinical Practice.

## Materials and Methods

We have prospectively screened all patients consecutively admitted between June the 14^th^ until August 11^th^ with confirmed *International Classification of Diseases* (ICD) 10 code of ‘U07.1 COVID-19 or ICD-10 code of ‘U07.2 COVID-19 diagnosis.

The study designed is presented in the *STrengthening the Reporting of OBservational studies in Epidemiology* (**STROBE**) diagram, **Figure 1**.

**Figure 1.**
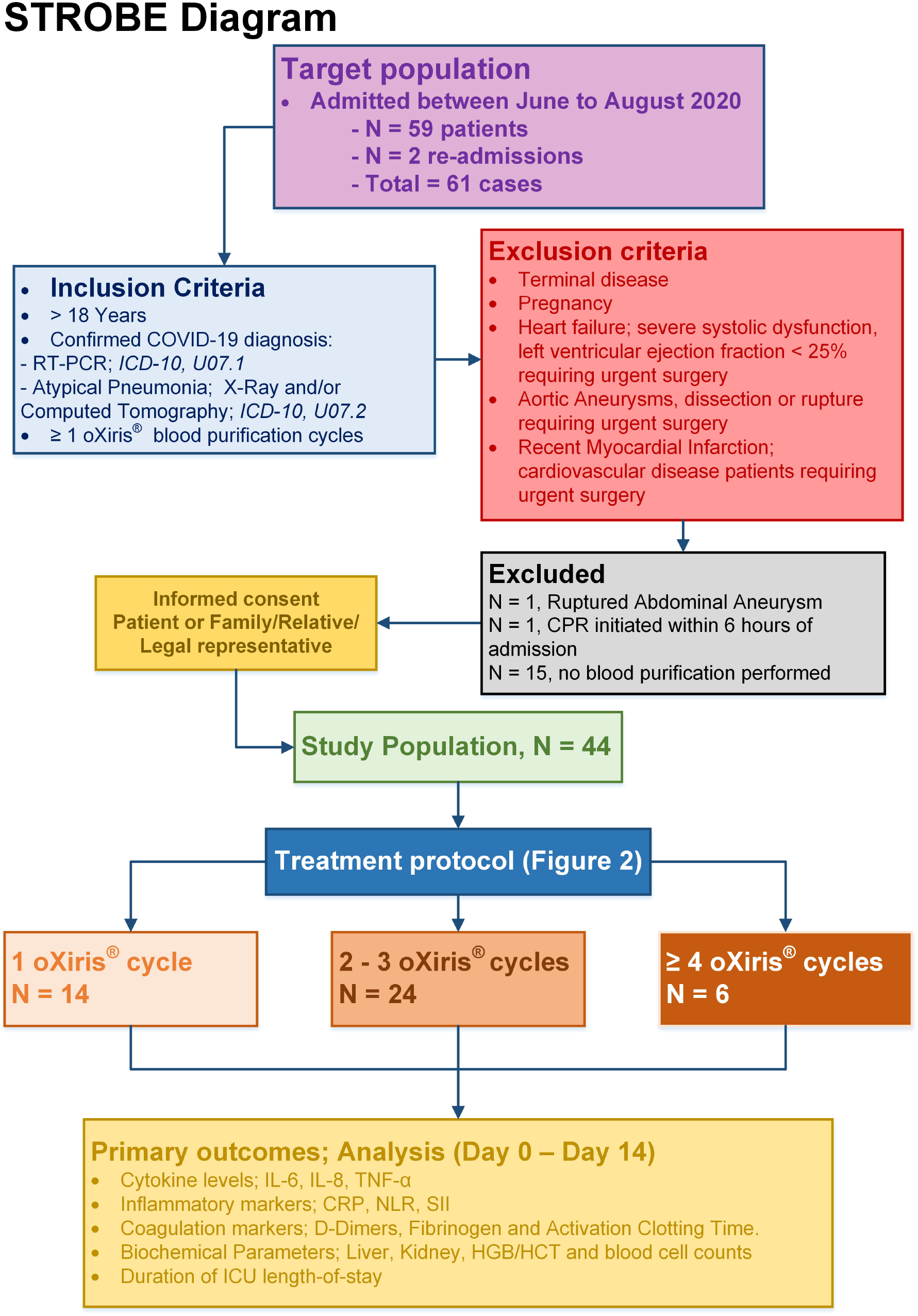
STrengthening the Reporting of OBservational studies in Epidemiology (STROBE) diagram.

Patients were classified according to their clinical presentation in 4 severity degrees:

1. **Mild cases** The clinical symptoms are mild, with no apparent sign of pneumonia on imaging.
2. **Moderate cases** Showing fever and respiratory symptoms with radiological findings of pneumonia.
3. **Severe cases**

A. Respiratory distress (30 breaths/min)
B. Oxygen saturation < 93% at rest *Cases with chest imaging that show lesion* > *50% progression within 24 hours shall be managed as severe cases*.
4. **Critical cases**

A. Respiratory failure requiring mechanical ventilation
B. Shock
C. With organ failure that requires ICU care.

**The inclusion criteria and exclusion criteria are presented in Figure 1**

### Biochemistry Analysis

Blood samples were collected from each patient at the time-points shown for routine blood analysis: White blood cell count (WBC), Lymphocyte count (LYM), Neutrophil count (NEU), Thrombocyte count (PLT), Monocyte count (MONO) and Eosinophil count (EO) were determined as well as the Neutrophil-to-Lymphocyte ratio (NEU/LYM) and the systemic immune-inflammation index PLT*(NEU/LYM). Moreover, blood biochemistry parameters such as Na^+^, K^+^ Aspartate aminotransferase (AST), Alanine aminotransferase (ALT), Urea, C-Reactive Protein (CRP), Interleukin-6 (IL-6) as well as Procalcitonin, lactate dehydrogenase (LDH), were assessed using Siemens ADVIA Centaur XP Immunoassay System.

Data on coagulation parameters were obtained from all patients; coagulation tests included D-dimers, Fibrinogen (FIB) and international normalised ratio (INR). Tests were performed using a Sysmex CA-600 automatic coagulation analyser. Blood Gas Analysis (BGA) was performed on a Siemens rapidpoint 500®.

### Statistical analysis

Categorical parameters were summarised as absolute numbers and percentages. Continuous data are shown as mean ± SD or median + Interquartile range (IQR).

Continuous variables were evaluated using the D’Agostino & Pearson normality test – Independent data that follow the Gaussian distribution were analysed via the *Student’s t-test*, and non-Gaussian continuous variables were assessed via the Mann-Whitney test for independent comparisons. Comparisons of preoperative vs postoperative data were performed using a Paired T-test or Wilcoxon Signed Rank test for non-parametric data.

The Fisher exact test was used to evaluate the association between categorical variables with the outcome. Regression and/or correlation analysis between the biomarkers over time were performed using Pearson’s or Spearman’s rank correlation testing.

The data were analysed with the statistical programs Graphpad Prism (version 8.43) and Statsdirect (version 3.3.3)

### Treatment

The general treatment protocol is shown in **Figure 2** and follows practice safety recommendations, treatment strategies and up-to-date sepsis management guidelines [18-21]. The multidisciplinary care and therapeutic approach were previously described in detail by Ugurov et al. [15]

**Figure 2.**
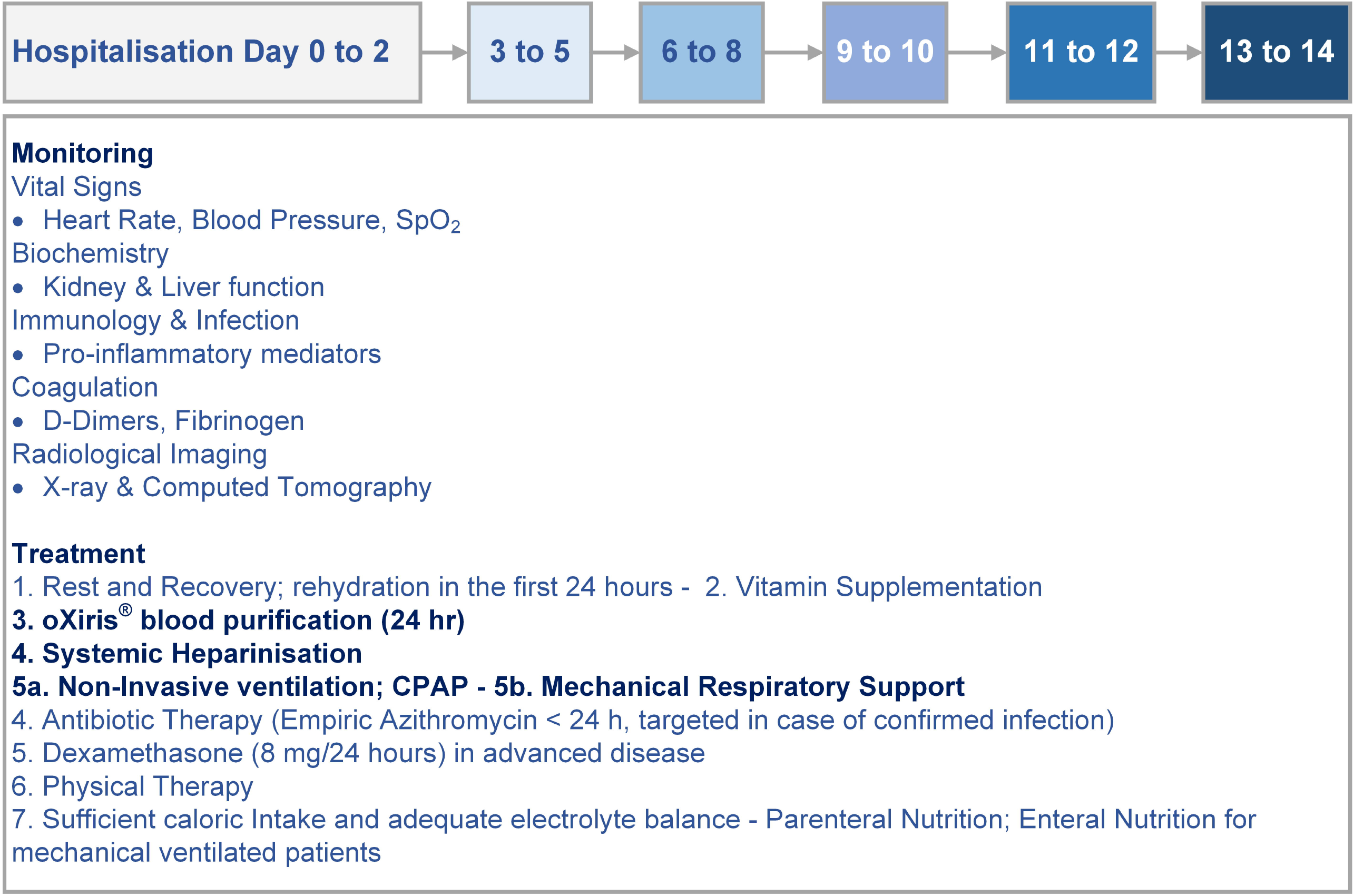
Treatment protocol for COVID-19 patients. In addition to blood purification, systemic heparinisation and physical therapy under continuous positive airway pressure (CPAP), the general care protocol consisted of rest and recovery; sufficient caloric intake and adequate electrolyte balance; aggressive rehydration in the first 24 hours; parenteral, enteral nutrition for mechanically ventilated patients; low-dose Dexamethasone therapy (8 mg/24 hours), antibiotic therapy, and biochemical and chest X-ray imaging for monitoring

### Extracorporeal Blood Purification

The Prismaflex® oXiris® system was mounted in the ICU and connected 4 - 12 hours after admission upon establishing control of the haemostasis, ACT = Activated Coagulation Time of 180 secs. The patient is connected to the Prismaflex® oXiris® system via a double lumen catheter placed in the femoral vein or vena subclavia.

## Results

### Patient demographics and clinical characteristics at admission

Among the 61 patients admitted with COVID-19 in the selected period, 42 were treated with oXiris® and thus considered in this single-centre case series. Of these, 2 patients were readmitted and counted as independent cases, totalling 44 consecutive cases, **Figure 1**.

The clinical characteristics and demographics are described in **Table 1**. At admission, 68.2% (N = 30) could be classified as severe, and 32.8% (N = 14) with mild to moderate symptoms (= non-severe). Primary symptoms reported were dyspnea, fever and breathing difficulties; all but one case (58/59, 98.3%) had confirmed COVID-19 pneumonia at admission **Supplemental Table 1 and 2**.

**Table 1.** Patient Characteristics, Demographics.

|  | Full cohort<br>N = 44 | Mild/Moderate<br>N = 14 | Severe<br>N = 30 | Effect Size<br>Mild/Moderate vs Severe |
| --- | --- | --- | --- | --- |
| Age (years) mean ± SD | 59.7 ± 12.0 | 53.9 ± 16.5 | 62.4 ± 8.4 | <sup>2</sup>8.5 [CI95% 0.9 to 15.9] $p=0.026$ |
| Age > 65 years, N (%) | 16 (36) | 4 (29) | 12 (40) | <sup>1</sup>1.7 [CI95% 0.4 to 5.6] $p=0.52$ |
| **Gender** |  |  |  |  |
| Male, N (%) | 36 (82) | 13 (93) | 23 (77) | <sup>1</sup>42.3 [CI95% 4.9 to 469] $p<0.0001$ |
| Female gender, N (%) | 8 (18) | 1 (7) | 7 (23) |  |
| Type 2 Diabetes, N (%) | 7 (16) | 0 (0) | 7 (23) | <sup>1</sup>∞ [CI95% 1.5 to infinity] $p=0.03$ |
| Hypertension, N (%) | 22 (53) | 6 (53) | 16 (53) | <sup>1</sup>5.3 [CI95% 1.0 to 20] $p=0.036$ |
| BMI | 27.6 ± 3.5 | 26.7 ± 3.3 | 29 ± 4.8 | <sup>1</sup>2.6 [CI95% -0.3 to 5.4] $p=0.081$ |
| Obesity*, N (%) | 13 (30) | 2 (14) | 11 (37) | <sup>*</sup>3.9 [CI95% 1.3 to 14.3] $p=0.03$ |
| SpO<sub>2</sub> (%) | 93.1 ± 5.8 | 94.7 ± 2.8 | 92.4 ± 6.6 | 9 [CI95% 6 to 15] $p<0.0001$ |
| Heart Rate (Beats/min) | 96.4 ± 22.1 | 79.2 ± 13.9 | 102.4 ± 21.3 | 23 [CI95% 21.3 to 24.9] $p<0.0001$ |
| Respiratory Rate (breaths/min) | 24 (20 - 29) | 22 (18 - 26) | 25 (21 - 29) | 3 [CI95% 3 to 4] $p<0.0001$ |
| mean Blood pressure (mmHg) | 127.4 ± 29.6 | 125.2 ± 16.4 | 127.9 ± 31.2 | 2.7 [CI95% -6.6 to 11.9] $p=0.201$ |
| Symptom duration (days) | 7 (5 - 7) | 5 (5 - 7) | 7 (6 - 8) | 2.0 [CI95% 0 to 3] $p=0.025$ |
| **Referral** |  |  |  |  |
| Home, N (%) | 19 (43) | 9 (64) | 10 (36) | <sup>*</sup>3.6 [CI95% 0.98 to 13.3] $p=0.101$ |
| peripheral clinic, N (%) | 25 (57) | 5 (36) | 20 (67) |  |

Median (Interquartile range)
Mean ± Standard Deviation
<sup>1</sup>median difference
<sup>2</sup>mean difference
\*BMI > 30 Kg/m<sup>2</sup>
<sup>\*</sup>Fisher exact test, odds ratio
ND = Not detected
NR = Not relevant

We have highlighted 3 unique and challenging cases in **Figure 3**.

**Figure 3.**
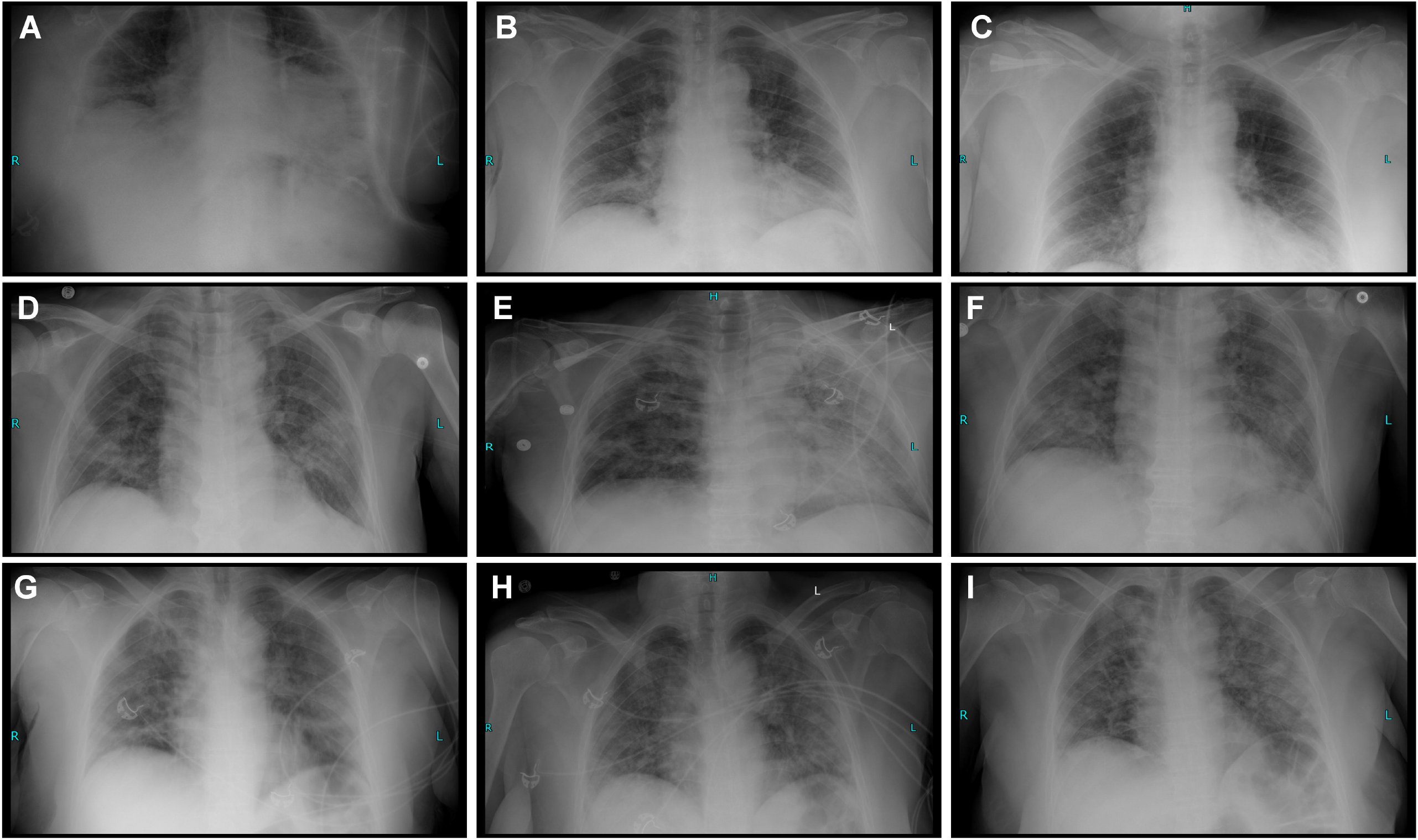
Radiography timeline and clinical course description of 3 COVID-19 cases treated with hemofiltration. **Upper panels**, a 71-year-old woman was admitted to our ICU with breathing difficulties for five days. Her peripheral oxygen saturation was 75% without oxygen support, and chest X-ray revealed diffuse atypical pneumonia (**A**). The laboratory findings showed suggested hyperinflammation and hypercoagulability, CRP, Fibrinogen, D-dimers and Ferritin of 208.9 mg/L, 5.5 g/L, 12980 ng/mL and 1198.4 ng/mL, respectively. Treatment with non-invasive ventilation was started and prone positioning. Continuous venovenous hemofiltration (CVVHF) using oXiris® filter was initiated in parallel. After two cycles of hemofiltration, the biochemical picture improved. The respiratory condition of the patient also improved (**B**). On day eight, after admission, the third cycle of hemofiltration was performed because of fever (38.3%) and a *de novo* increase of inflammatory markers. Afterwards, the patient recovered well and was discharged on day 18th after admission with 1-2L O_2_ support SpO_2_ 91-93% and improvements on the lung pictures (**C**). The clinical improvement of this patient was also associated with remarkable normalisation of eosinopenia, from 0.14% at admission to 5.6% at discharge. **Middle panels**, a 64-year-old man, was admitted to our ICU from home with fever (38.4°C), fatigue and cough. He was diagnosed COVID-19 three days earlier. Chest X-ray (**D**) showed bilateral atypical inflammation, and the laboratory findings showed high levels of inflammatory markers during our care (peak levels of CRP, IL-6, Ferritin, LDH, Fibrinogen and D-dimers were 177.9 mg/L, 142 pg/mL, 1347 ng/mL, 586 U/L, 8 g/L and 7791 ng/mL, respectively. Continuous venovenous hemofiltration using oXiris® filter was initiated within 8 hours after admission and performed a total of 5x cycles. On day five after admission, the patient required oxygen support (SpO_2_ 89%, 4L O_2_). The patient was treated with non-invasive ventilation and was placed in a prone position. Chest X-ray showed significant bilateral progression of inflammation in the lungs, in particular on the left lobe (**E**). Because of repeating episodes of fever (Procalcitonin 0.21 ng/mL), a broad-spectrum antibiotic was started to counter a possible superinfection. The patient underwent five cycles of continuous venovenous hemofiltration using oXiris® filter. On day 14 after admission, the patient required no oxygen support; his clinical condition was gradually improving, and we were able to discharge the patient after 21 days; radiography pointed to residual lung injury and signs of fibrosis (**F**). Follow up after 30 days pointed to further improvement according to biomarkers (Fibrinogen 8.0 g/L vs 5.5 g/L at discharge, D-dimers, 1930 ng/mL vs 6250 ng/mL at discharge and LDH 173 U/L vs 586 U/L. However, he reported signs of cardiovascular disease (NYHA III); the patient will remain under continuous monitoring. **Lower panels**, 57-year-old women were transferred to our ICU from another peripheral hospital because of worsening of oxygen saturation (SpO_2_ 75%, 6-8 L O_2_, respiratory rate 35/min). Chest X-ray showed bilateral atypical inflammation (**G**). Diagnosis via radiography and RT-PCR was accompanied by high levels of Lactate Dehydrogenase (453 U/L), C-reactive protein (299.5 mg/L) and Neutrophilia (90.2%). CVVHF using Oxiris filter (Prismaflex, Gambro). In total, three cycles were performed with a duration period of 24 hours per cycle. Simultaneously the patient was treated with non-invasive ventilation and was placed on prone position. The sputum culture showed Escherichia coli and Candida species. Escherichia coli was treated with Ceftriaxone and Candida species with fluconazole. On day seven after admission, the patient had 93% SpO_2_ saturation with nasal flow cannula 2l/min (**H**). The patient was discharged on day 11 with no need for oxygen support but with significant lung fibrosis (**I**).

Patients with severe disease were characterised by more prolonged duration of symptoms; they were admitted on average 7 days (vs 5 days for non-severe cases, *p=0*.*025*) after symptom onset and were more likely to be referred from peripheral clinics (67% vs 36%, OR 3.6 [CI95% 0.98 to 13.3], *p = 0*.*101)* in comparison to direct admission following self-isolation at home.

We observed non-significant trends suggesting more pronounced respiratory difficulties and lower blood pressure in advanced disease. We aggregated selected laboratory values and patient signs that accounts for both acute and chronic disease to determine the Acute Physiology And Chronic Health Evaluation II (APACHE II) at admission; severe cases had an overall APACHE II score of 8.5 ± 4.6 vs 4.7 ± 3.0, mean difference 3.5 [CI95% 3.5 to 6.1], *p=0*.*011*.

Severe cases were admitted with a more prominent inflammatory profile, demonstrated by elevated levels of LDH, 494.5 U/L (IQR374 - 669); Ferritin, 1100 ng/mL (IQR406 - 1650); CRP, 151 mg/L (IQR82 - 211) and the immune indices NLR and SII, 11.5 (7.5 - 18.5) and 2580 (1554 - 4692), respectively. (**Table 2**). Moreover, we observed substantially higher D-dimers levels in severe cases compared to those with mild to moderate symptoms, median difference, 630 [CI95% 160.0 to 6220], *p=0*.*013*, (**Table 2**).

**Table 2.** Patient clinical data @admission.

|  | Full cohort<br>N = 44 | Mild/Moderate<br>N = 14 | Severe<br>N = 30 | Effect Size<br>Mild/Moderate vs Severe |
| --- | --- | --- | --- | --- |
| ALT (U/L) | 57 (30 - 85) | 55 (25 - 76) | 58 (31 - 107) | <sup>1</sup>3.5 [CI95% -13 to 36] $p=0.327$ |
| AST (U/L) | 42 (29 - 66) | 41 (26 - 66) | 44 (32 - 67) | <sup>1</sup>3 [CI95% -10.0 to 20.0] $p=0.528$ |
| Urea (mmol/L) | 5.6 (3.9 - 8.1) | 3.9 (3.4 - 5.9) | 6.7 (4.9 - 9.3) | <sup>1</sup>2.7 [CI95% 0.9 to 4.3] $p=0.0013$ |
| Creatinine (μmol/L) | 91.8 ± 32.2 | 81.9 ± 15.2 | 96.4 ± 36.9 | <sup>2</sup>13.4 [CI95% -6.3 to 35.5] $p=0.17$ |
| GFR (mL/min) | 108.3 ± 39.7 | 104.8 ± 42.6 | 115.6 ± 33.2 | <sup>2</sup>-10.8 [CI95% -37.0 to 15.4] $p=0.410$ |
| LDH (U/L) | 407 (272 - 569) | 252 (203 - 303) | 495 (374 - 669) | <sup>1</sup>243 [CI95% 142 to 358] $p<0.0001$ |
| RBC (*10<sup>6</sup> counts/μL) | 4.8 ± 0.7 | 4.8 ± 0.4 | 4.8 ± 0.8 | <sup>1</sup>0.02 [CI95% -0.46 to 0.50] $p=0.929$ |
| HCT (%) | 39.6 ± 5.8 | 39.5 ± 2.7 | 39.7 ± 6.9 | <sup>2</sup>0.11 [CI95% -3.8 to 3.9] $p=0.955$ |
| HGB (g/dL) | 13.8 ± 2.1 | 13.8 ± 0.9 | 13.8 ± 2.5 | <sup>2</sup>-0.01 [CI95% -1.4 to 1.4] $p=0.994$ |
| WBC (*10<sup>3</sup> counts/μL) | 7.4 (4.8 - 12.1) | 4.0 (2.7 - 6.5) | 9.8 (7.1 - 13.4) | <sup>1</sup>5.8 [CI95% 2.6 to 7.8] $p<0.0001$ |
| Basophils (%) | 0.14 (0.1 - 0.24) | 0.27 (0.2 - 0.4) | 0.14 (0.1 - 0.14) | <sup>1</sup>-0.1 [CI95% -0.3 to -0.1] $p=0.0003$ |
| Eosinophils (%) | 0.17 (0.03 - 0.47) | 0.12 (0.0 - 0.49) | 0.10 (0.04 - 0.24) | <sup>1</sup>-0.02 [CI95% -0.2 to 0.1] $p=0.97$ |
| Monocytes (%) | 4.0 (3.0 - 6.3) | 5.5 (4.0 - 11.7) | 3.6 (2.4 - 5.3) | <sup>1</sup>-1.9 [CI95% -5.9 to -0.8] $p=0.0017$ |
| Neutrophils (%) | 82.2 ± 11.4 | 72.3 ± 10.3 | 86.9 ± 8.5 | <sup>2</sup>14.7 [CI95% 8.7 to 20.7] $p<0.0001$ |
| Lymphocytes (%) | 9.9 (5.5 -13.4) | 20.1 (11.1 - 26.7) | 7.7 (4.9 - 11.2) | <sup>1</sup>-12.4 [CI95% -17.6 to -5.4] $p<0.0001$ |
| Platelets (*10<sup>3</sup> counts/μl) | 200 (127 - 256) | 138 (119 - 176) | 224 (146 - 337) | <sup>1</sup>62 [CI95% 13 to 142] $p=0.012$ |
| NLR (*10<sup>3</sup> counts/μl) | 8.5 (5.9 - 16.0) | 3.6 (2.5 - 7.6) | 11.5 (7.5 - 18.5) | <sup>1</sup>7.9 [CI95% 4.4 to 12.6] $p<0.0001$ |
| SII | 2047 (815 - 3594) | 620 (321 - 1071) | 2580 (1554 - 4692) | <sup>1</sup>1959 [CI95% 1224 to 3215] $p<0.0001$ |
| Interleukin-6 (pg/mL) | 15.5 (7.4 - 47.3) | 16.9 (5.9 - 33.1) | 14.7 (7.9 - 64.3) | <sup>1</sup>-2.3 [CI95% -9.3 to 38.2] $p=0.39$ |
| C-Reactive Protein (mg/L) | 109.0 (57.8 - 173.1) | 71.1 (8.9 - 109.7) | 150.6 (81.6 - 210.6) | <sup>1</sup>79.5 [CI95% 33.4 to 135.9] $p=0.001$ |
| Ferritin (ng/mL) | 948 (352 - 1573) | 479 (268 - 1155) | 1100 (406 - 1650) | <sup>1</sup>621 [CI95% 113 to 872] $p=0.1548$ |
| Procalcitonin (ng/mL) | 0.07 (0.05 - 0.13) | 0.05 (0.01 - 0.05) | 0.11 (0.07 - 0.19) | <sup>1</sup>0.08 [CI95% 0.03 to 0.11] $p=0.0002$ |
| D-Dimers (ng/mL) | 735 (398 - 2728) | 595.0 (292.5 - 850) | 1225 (595.0 - 11503) | <sup>1</sup>535 [CI95% 110 to 2860] $p=0.0108$ |
| aPTT (sec) | 22.4 (20.6 - 24.8) | 23.6 (21.9 - 25.6) | 21.8 (20.4 - 23.7) | <sup>1</sup>-1.8 [CI95% -3.6 to 0.3] $p=0.086$ |
| Fibrinogen (g/L) | 4.9 (4.1 - 7.0) | 4.5 (3.9 - 6.2) | 5.1 (4.2 - 8.0) | <sup>1</sup>0.55 [CI95% -0.5 to 1.4] $p=0.352$ |

Median (Interquartile range)
Mean ± Standard Deviation
<sup>1</sup>median difference
<sup>2</sup>mean difference

Despite the clinically significant increase of two key acute phase proteins and inflammatory markers, there was only a marginal increase in IL-6 across the cohort, 15.5 (7.4 - 47.3) pg/mL

### Extracorporeal blood purification; treatment approach and clinical practice

A total of 14 mild/moderate (56%) and 30 severe patients (88.8%) were suspected of hyperinflammation (= CRP ≥ 100 mg/L and/or IL-6 ≥ 40 pg/mL and/or Ferritin ≥ 500 ng/mL) and indicated for blood purification using the AN69ST (oXiris®) hemofilter in an attempt to control disease progression (**Supplemental Figure 1**).

Flow rates were maintained as follow; effluent dose 35 mL/Kg/h, dialysate 14 – 16 mL/Kg/h, blood 150 mL/min, replacement 16 −18 mL/Kg/h; patient fluid removal is tailored to the individual’s volume status, ≈ 100 - 250 mL/h. The oXiris^®^ CRRT modality was chosen according to the patient’s kidney function; continuous venovenous hemofiltration (CVVH), continuous venovenous hemodiafiltration (CVVHDF) or slow continuous ultrafiltration (SCUF).

The average levels of urea, creatine and the Cockcroft Gault Glomerular Filtration Rate (GFR) were 5.6 (3.9 - 8.1) mmol/L, 91.8 ± 32.2 µmol/L and 108.3 ± 39.7 mL/min, respectively. We observed 2 cases of significant kidney dysfunction (= GFR < 50 mL/min) in our COVID-19 cohort.

Consequently, CVVHF (42/44, 95.5%) was the predominant CRRT modality indicated with the primary aim to reduce inflammatory mediators. Alternatively, CVVHDF was used for renal support. Total run time on the Prismaflex system was 3303.8hr; on average severe cases were treated with 2 (IQR1 - 3) blood purification cycles, one more than the non-severe patients. The average duration of blood purification was 30.3 hours (20.8 - 44.9) (**Table 3**).

**Table 3.**
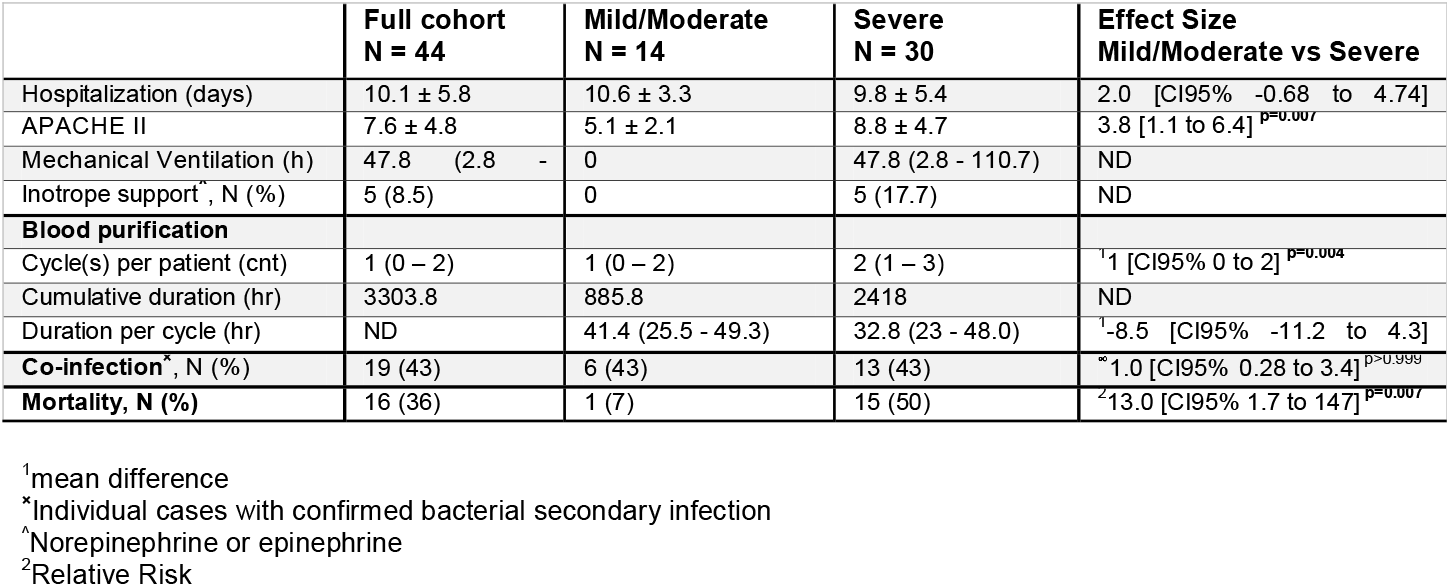
Clinical course and outcome.

We observed no complications related to bleeding or thromboembolism. There were 9 occurrences of premature clotting (machine run time < 5hr) resulting in a linearised incidence premature clotting rate of 0.27% (patient-hours).

One patient experienced a hematoma following venous cannulation that required a catheter displacement. Our complete treatment approach was previously documented [15]

In parallel, during hospitalisation, 12 critical patients were treated with i.v. Dexamethasone at a dose of 8 mg q.d.

### Time-series analyses of inflammatory biomarkers, vital signs and blood gas analysis parameters

Blood purification was associated with gradual normalization of several biomarkerslinked to COVID-19 disease severity (**Figure 4**). We observed a sharper reduction of Ferritin in severe pateints (**Fig 4A**), β = −23.0 [CI95% −36.79 to −9.251], *p = 0*.*0012, vs* β = −1.88 [CI95% −20.67 to 16.90], *p = 0*.*841* for the sub-group with milder symptoms).

**Figure 4.**
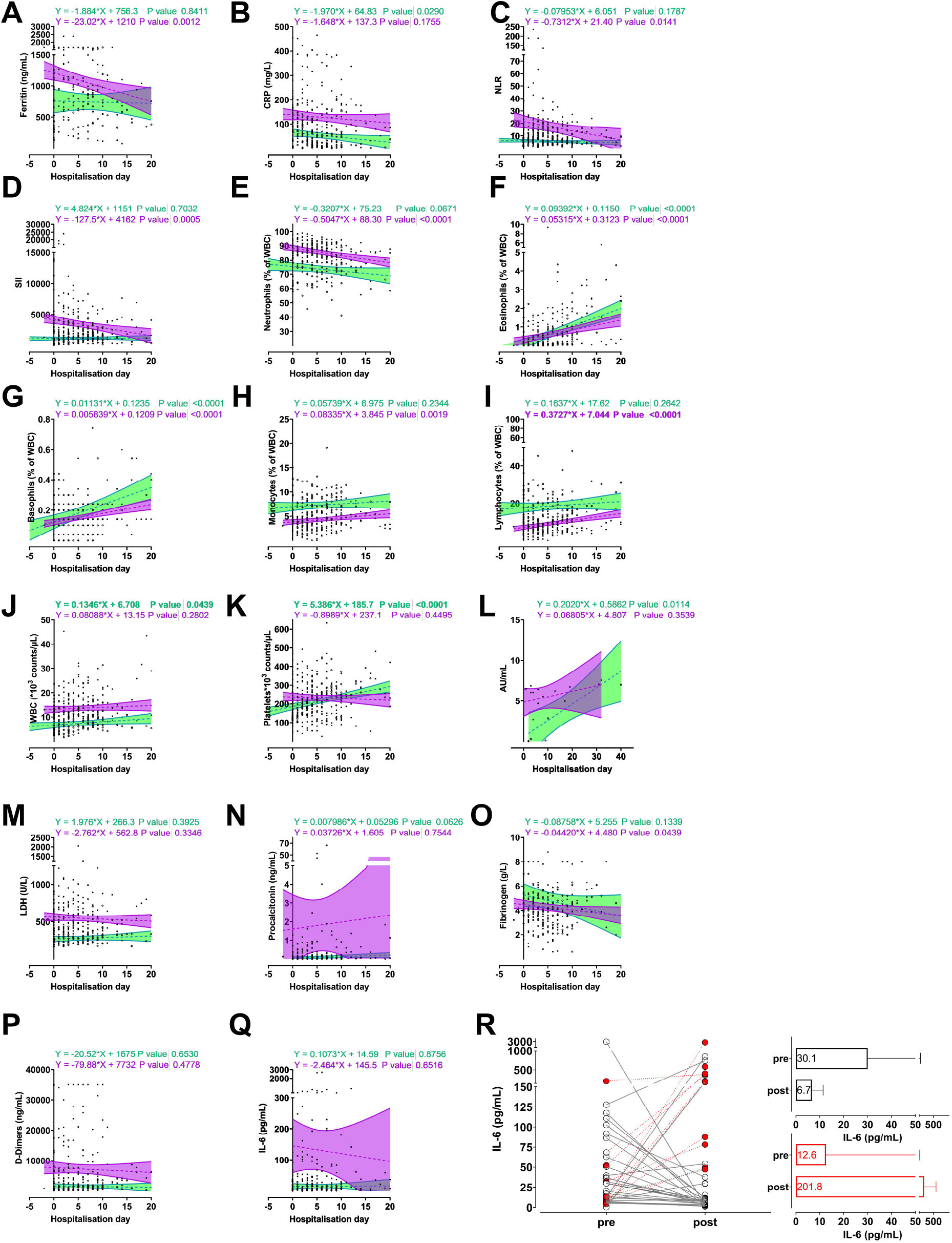
Longitudinal biomarker profiling of severe COVID-19 patients treated with extracorporeal blood purification. Graphs (**A – Q**) display concentrations (Ferritin, CRP, Procalcitonin, Fibrinogen, D-dimers and IL-6), percentages (Neutrophils, Eosinophils, Basophils, Monocytes, Lymphocytes), absolute counts (White Blood Cells and Platelets), and units (SARS-CoV-2 Immunoglobulin G, Lactate Dehydrogenase) over time stratified according to the treatment regimen; **purple** trendline indicates the severe sub-group vs **green** trendline for the subgroup with mild-to-moderate symptoms. Equations on the top of the graphs represent the respective regression trends for each treatment group during hospitalisation + significance level. Panel **R** represents individual IL-6 values before (pre) and after (post) blood purification using oXiris® hemofilter; adjacent graphs show the aggregated IL-6 data for hemofilter-treated COVID-19 patients who were successfully discharged (**upper bar graphs**), pre- vs post blood purification median difference of −21.44 pg/ml [CI95% −45.60 to −4.100], p=0.0001. Mortality cases (**lower bar graphs**), median difference 179.7 pg/ml [CI95% 42.02 to 613.3], p=0.0009.

Likewise, EBP was associated with a decrease of CRP with treatment in non-severe cohort showing a significant time-dependent reduction β = −1.9 [CI95% −3.7 to −0.21], *p = 0*.*03* (**Fig 4B**).

Moreover, EBP was also characterized by a decrease of inflammatory immune indices NLR, β = −0.73 [CI95% −1.31 to −0.15], *p = 0*.*01* (**Fig 4C**) and SII, β = −72.3 [CI95% −30.2 to −6.0], *p = 0*.*004*, (**Fig 4D**). Neutrophilia (>75% neutrophils of total white blood cells) was a notable feature of our cohort (**Table 2**), blood purification was associated with a significant decline in peripheral neutrophils (**Fig 4E**), and was also paired with the resolution of several cytopenias. Baseline levels of eosinophils, basophils, monocytes, lymphocytes in severe patients normalised during hospitalisation leading to increased numbers of WBC (**Table 2 & Fig 4F-J**).

Non-severe cases had lower baseline thrombocyte counts, 138*10^3^ counts/µL (IQR119 - 176) compared to severe patients, median difference −62*10^3^ counts/µL [CI95% 13 to 142], *p=0*.*012*, which was normalised within a week (**Fig 4K**).

We did not routinely perform screening for SARS-CoV-2 spike protein-specific IgG antibodies at admission; nevertheless, the available data suggests that severe cases presented with higher baseline titers which further increased time (**Fig 4L**).

Levels of LDH were discriminant at admission (**Table 2**), and the differences between subgroups persisted despite EBP (**Fig 4M**). Analysis of procalcitonin levels yielded similar observations (**Table 2, Fig 4N**).

Hypercoagulability state was assessed via Fibrinogen and D-dimers; we noted a slight decrease in Fibrinogen levels over time in non-severe cases (**Fig 4O**). On the other hand, D-dimers remained significantly elevated despite hemofiltration (**Fig 4P**).

IL-6 is the primary inducer of hepatic CRP synthesis and secretion; individual analysis indicates a periprocedural decrease of both IL-6 and CRP (**Supplemental figure 1**). Nonetheless, group analyses suggest that the time-dependent decrease in IL-6 was not significant (**Fig 4Q**).

Paired analysis may explain the mixed response of IL-6 to blood purification; clinical recovery was associated with a decrease of IL-6 levels, median difference −21.44 pg/ml [CI95% −45.6 to −4.1], *p=0*.*0001*. Whereas IL-6 continued to rise in patients who ultimately succumbed to the disease despite repetitive hemofiltration cycles (**Fig 4R**); median difference 179.7 pg/ml [CI95% 42.02 to 613.3], *p=0*.*0009*.

Finally, haematological parameters showed a continuous declining trend (**Figure 5**). Liver enzymes remained stable during hospitalisation; however, whereas creatine remained within normal ranges, there was a rise of blood urea evident in the severe cohort (**Fig 5F**).

**Figure 5.**
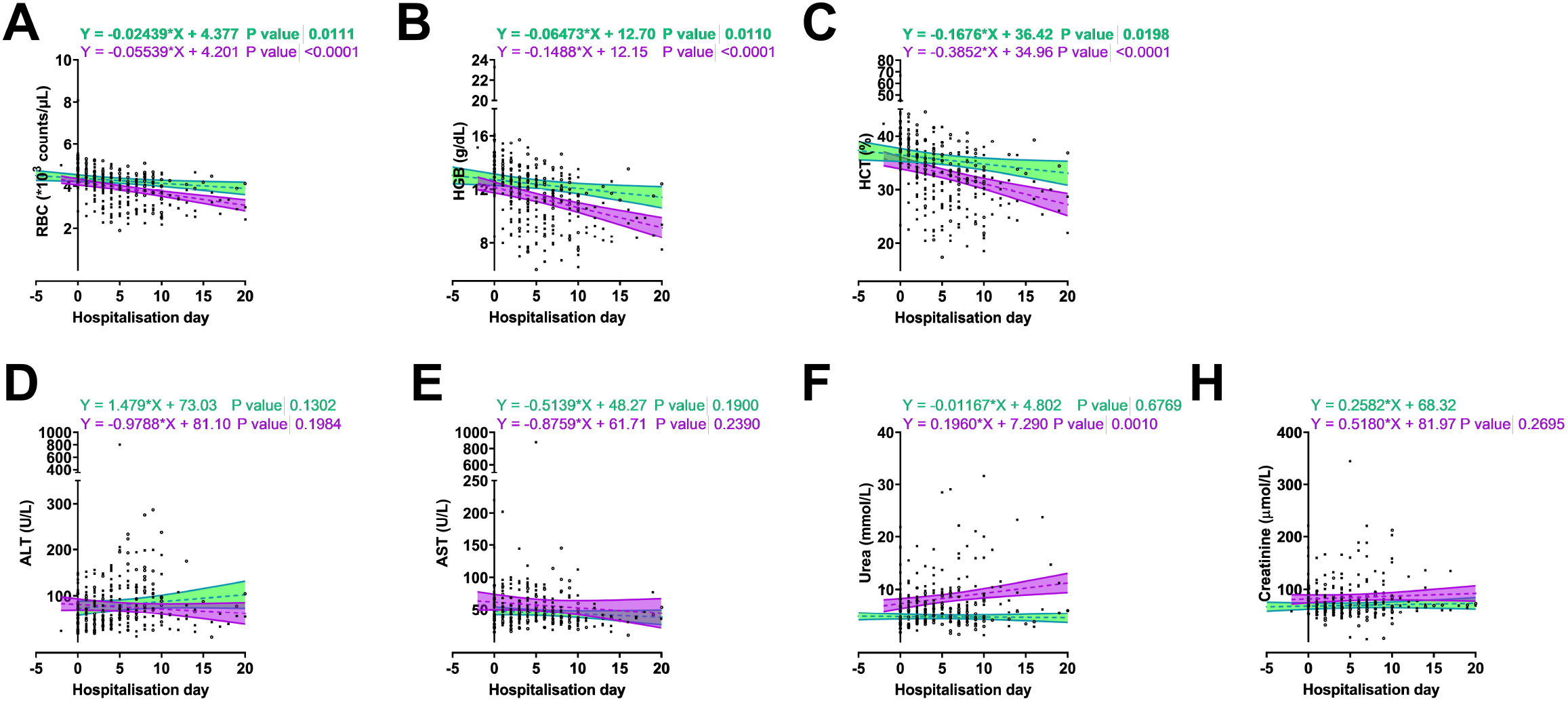
Longitudinal Kidney and Liver function and haematological parameters of COVID-19 patients treated with extracorporeal blood purification. Graphs display systemic concentrations of creatinine, blood urea, aspartate transaminase (AST) and Alanine transaminase (ALT), Red Blood Cells (RBC), Hematocrit (HCT), Hemoglobulin (HGB) over time stratified according to the treatment regimen; **purple** trendline indicates the severe sub-group vs **green** trendline for the subgroup with mild-to-moderate symptoms. Equations on the top of the graphs represent the respective regression trends for each treatment group during hospitalisation + significance level.

Clinically, both subgroups were characterised by normalising vital parameters over time (**Figure 6**); in particular, severe patients experienced a gradual improvement in SpO_2_ and breathing capacity. Blood gas analyses (BGA) were comparable during the early phases of hospitalisation; excluding the individual exceptions, there were no notable differences in BGA parameters over time across the cohort.

**Figure 6.**
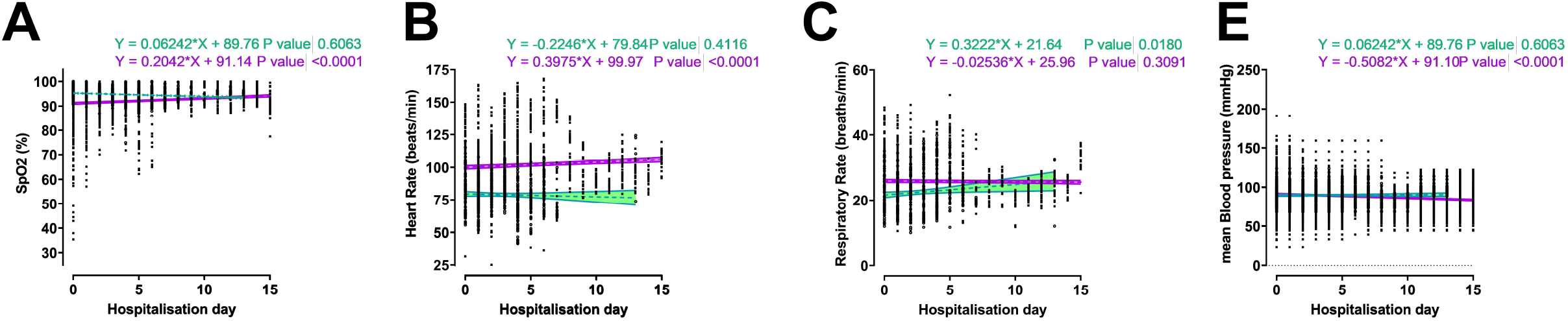
Timeseries analyses of COVID-19 patients treated with extracorporeal blood purification. Graphs display systemic concentrations of Blood Gas and related markers (pH, pCO_2_, pO_2_ and Base Excess), the electrolytes Na^+^, K^+^, and Ca^2+^ and the metabolites, Glucose and Lactate; **purple** trendline indicates the severe sub-group vs **green** trendline for the subgroup with mild-to-moderate symptoms. Equations on the top of the graphs represent the respective regression trends for each treatment group during hospitalisation + significance level.

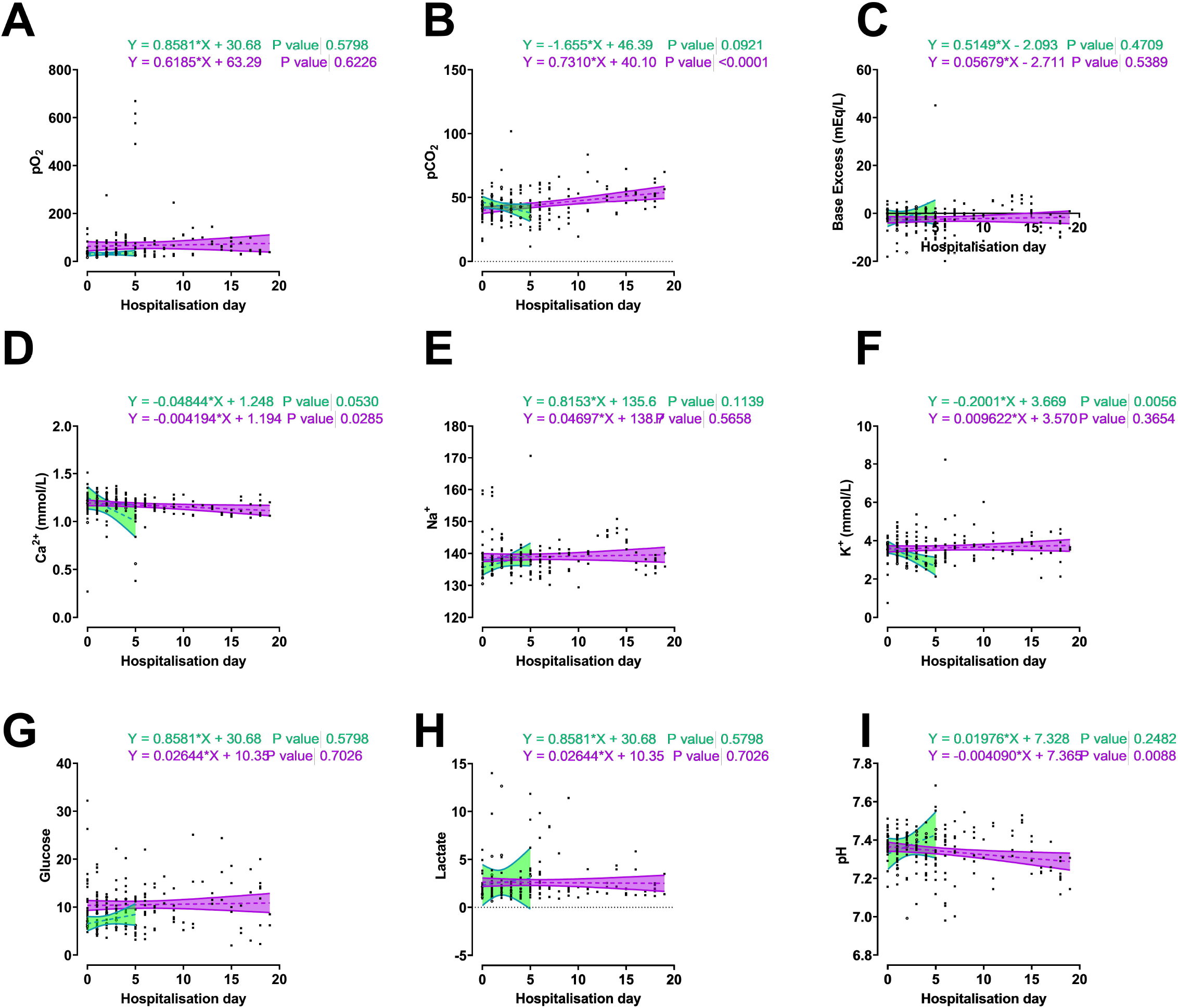

Severe cases requiring prolonged hospitalisation (> 2 weeks) showed possible signs of respiratory acidosis (ARDS) evidenced by decreasing pH-values and increasing levels of the partial pressure of carbon dioxide (pCO_2_) on mechanical circulatory support.

We observed 19 cases (43.2%) (**Table 3**) of secondary bacterial co-infections, most confirmed microbiology cultures pertained to samples collected at admission; furthermore, positive microbiology was most likely to occur in patients admitted from peripheral clinics, OR 3.6 [CI95% 0.9 to 12.4], *p=0*.*07*. We most frequently detected *Staphylococcus aureus, Haemophilus influenzae*, and Enterobacteriaceae; however, we also detected *Klebsiella pneumoniae* and *Acinetobacter baumannii*. The mortality rate over the whole cohort was 27%; the rate in severe cases was 44% vs 4% in non-severe, relative risk (RR) 7 [CI95%1.49 to 40.2], *p=0*.*007*.

## Discussion

COVID-19 is caused by the severe acute respiratory syndrome coronavirus 2 (SARS-CoV-2) and was identified by the World Health Organization (WHO) as an international public health emergency; with that, a multitude of therapeutic approaches have been pioneered to counter the detrimental clinical manifestations [22-25] linked to inflammatory immune disturbances

We here describe the clinical course, outcome and longitudinal analysis of multiple biomarkers, clinical and blood gas parameters in non-severe vs severe COVID-19 patients treated with blood purification using the cytokine adsorbing oXiris® hemodiafilter.

We and others have explored the use of cytokine blood purification in an attempt to prevent, control or reduce (suspected) hyperinflammation [26-30]. Several blood-purification devices are currently on the market, showing a significant reduction of pro-inflammatory cytokines and restore homeostatic dysregulation associated with systemic inflammatory syndromes [31-33].

The United States Food and Drug Administration (FDA) gave emergency approval to the oXiris® membrane filter and CytoSorb® column-based filter. Both devices have shown an exceptional capacity to reduce cytokine levels [32, 34], support hemodynamic stabilisation and increase survival probability, especially in patients whom therapy was initiated early [35, 36].

We previously reported our preliminary results on EBP using the oXiris® filter in COVID-19 [15]. The cohort’s limited size precluded comparative analyses between patient subgroups stratified according to disease severity. With this follow up analysis on an expanded cohort we show that EBP was paired with longitudinal reduction of numerous inflammatory mediators, acute phase proteins and resolution of cytopenias (**Figure 3**).

For instance, EBP was associated with a time-dependent decrease across the cohort in Ferritin, a well-described biomarker associated with severe COVID-19 [37, 38]. Interestingly, oXiris® was associated with significant decreases over time of CRP in non-severe cases but not in severe cases.

Moreover, our results suggest a weaker association between IL-6 and CRP in COVID-19 patients treated with EBP; the two markers showed a weak(er) correlation (r = 0.05, *p =* 0.66, non-severe and r, = 0.35, *p*<*0*.*001* severe cases (**Supplemental Figure 2**). IL-6 is the main stimulus for CRP synthesis and others have described a strong positive correlation between IL-6 and CRP in numerous disease conditions [39-41].

Systemic levels of CRP were more detected in critical ranges compared to IL-6. The cytokine was increased in severe cases, but overall, the systemic levels detected were lower than observed in our historical bacterial sepsis cohort.

The higher levels of CRP irrespective of IL-6 might be explained as follows: IL-6 is the primary driver of increased production of CRP; however, during injury or other trauma, other pro-inflammatory cytokines such as IL-1, TNF-α and IL-8 may contribute the production of CRP [42-44].

Current literature indicates that although IL-6 is essential for CRP gene induction, it is not sufficient to achieve this alone [45] and might require orchestrated stimuli by multiple pro-inflammatory cytokines to boost CRP gene expression. Collectively, the increased levels of IL-1, TNF-α, IL-8 and IL-6 seen in COVID-19 plausibly synergise to enhance CRP gene expression [46].

Recently, three studies have compared cytokine levels in COVID-19 to other conditions [46-48] and hinted that the term “cytokine storm” is perhaps not applicable to COVID-19 as was previously postulated.

Possibly a case of semantics; the immunological disturbances in COVID-19 encompass both innate and adaptive arms [46, 4, 49] and clinically possess many hallmarks of cytokine storm syndrome [50, 51].

In our experience, abnormal neutrophil counts and increased neutrophil-to-lymphocyte ratios (NLR) often accompanied hyperinflammation. Neutrophilia was shown to coincide with lung lesion injury on CT [52] and predict poor outcomes in COVID-19 patients [8]. The NLR is also an independent risk factor for adverse outcome [53, 52]. Our results point to higher NLR in severe patients and EBP was associated with normalizing neutrophil counts and NLR.

Along similar lines, LDH is strongly linked with disease severity; elevated LDH serum concentrations reflect tissue/cell destruction and are regarded as a common sign of tissue/cell damage [54]. Serum LDH has been identified as an important biomarker for the activity and severity of idiopathic pulmonary fibrosis [55].

For critically ill patients with COVID-19, the rise in LDH level may indicate an increase of the activity and extent of lung injury. To this end, LDH has emerged as a prominent variable to distinguish the majority of COVID-19 patients who require immediate medical attention [56]. In our study, the patients that succumbed to COVID-19 all showed repetitive measurements of LDH surpassing the critical threshold of 365 U/L linked to mortality [56].

The incidence of bacterial co-infection (43%) was higher than previously reported [57-59]. In compliance with antimicrobial resistance (AMR) guidelines [60, 61], we collect nasopharyngeal samples and also perform hemocultures in critical COVID-19 cases at admission. We detected bacterial co-infections more frequently in patients previously hospitalised at peripheral clinics compared to the sub-group admitted from home.

Empirical antibiotics are administered for max 48hr or discontinued immediately following negative representative cultures [62]. It is possible that our AMR-protocol may be masked by unwarranted antibiotics administration elsewhere, a common clinical practice in south-eastern Europe [63-66].

Clinically, the heterogeneity among the severe cases was striking; the clinical course of severe COVID-19, the extent of the affected lung area and development of ARDS was often not aligned with systemic levels of biomarkers. Several critical cases in need of mechanical ventilation and inotrope support showed clinical deterioration despite decreasing inflammatory mediators.

A consistent observation in our study was the gradual decrease in SpO_2_ in critical cases and the continuous high values of D-dimers, which may point to hypercoagulability, micro-embolism, leading to inadequate peripheral oxygen homeostasis.

Interestingly, we observed just two cases of AKI during the study period; a surprising finding given that the pooled incidence rate of 8.9% (CI95% 4.6 - 14.5) reported by Chen et al. [67]

Finally, the observed mortality rate of 27% observed in this case series is comparable to what has been recently reported [68].

### Limitations

Because of the observational design, absence of randomisation and limited cohort size are unable to provide concrete clinical guidance on the optimal EBP strategy to control disease progression and prevent ARDS. Nevertheless, with 3000+ accumulated hours of EBP run-time and real-time data on 44 patients we collected sufficient datapoints to establish biomarker fluctuations over time and evaluate the safety-profile of oXiris® blood purification.

## Conclusion

Our study results indicate that extracorporeal blood purification with oXiris® was associated with reduced levels of acute-phase proteins, resolution of cytopenias and control of hyperinflammation.

EBP represents an attractive treatment modality to limit systemic damage caused by aberrant immune activation and therefore may stabilise the clinical condition of COVID-19 patients.

## Data Availability

The data used for this study are currently in transfer in the SCCM COVID-19 registry, https://sccmcovid19.org/. Until upload is completed the data is available for review upon reasonable request by editorial staff members.

## List of abbreviations

IL: interleukin
TNF: tumor necrosis factor
CRP: C-reactive protein
ARDS: acute respiratory distress syndrome
AKI: Acute Kidney Injury
CPAP: Continuous Positive Airway Pressure
EBP: Extracorporeal Blood Purification
CVVH: continuous venovenous hemofiltration
CVVHDF: continuous venovenous hemodiafiltration
SCUF: slow continuous ultrafiltration
NIV: Non-invasive ventilation

## Statements

## Acknowledgements

None

## Statement of Ethics

The local ethical committee of the Zan Mitrev Clinic reviewed and approved the clinical practice, treatment procedures described, and the results reported in this manuscript and approved the submission, #EBPZ.357. Trial registration: ClinicalTrials.gov, NCT04478539. Registered 14^th^ of July 2020 - Retrospectively registered, https://clinicaltrials.gov/ct2/show/NCT04478539

Written informed (or temporary verbal) consent was obtained from all patients for publication of this manuscript and any accompanying images; the use of all health and medical information for scientific research and manuscript preparation was approved. A copy of the written consent is available for review by the Editor-in-Chief of this journal. For those patients that were not traceable after discharge, the need for informed consent was waived by the ethical committee.

## Conflict of Interest Statement

Dr Zan Mitrev is the hospital director at the *Zan Mitrev Clinic*. Gianluca Villa has received support for travel expenses, hotel accommodations and registration to meetings from Baxter. The other co-authors do not have any competing interests to disclose.

## Funding Sources

Not applicable

## Authors’ contributions

ZM is the study director; PU, DP, DN, LVK and ZM were responsible for diagnostics and patient care. L.V-K. performed the radiological examinations. RR coordinated the study, provided academic assistance, managed data collection, analysed the data and wrote the manuscript with the assistance of PU GV provided academic input and writing support and critically reviewed the manuscript. AT and DK provided IT-support for data validation/extraction. All authors have read and approved the manuscript.

## Figure Legends

**Supplemental Figure 1 - Inflammatory mediator analysis; systemic levels of IL-6 and CRP during hospitalisation**

Representative individual profiles showing time-series data on IL-6 (pg/mL) and C-reactive protein (CRP) (mg/L) are plotted on the left y-axis. The start of oXiris^®^ Hemofiltration 24-cycle is depicted as a blue checkered line on the x-axis; day 0 is defined as the first 24 hours following admission. One patient also received Tocilizumab (= anti-IL6 receptor mAb) indicated in a green checkered line.

**Supplemental Figure 2 - Biomarker correlation in COVID-19 patients treated with extracorporeal blood purification**.

Correlation matrix depicts Pearson’s *r*, and colour gradient visualise the degree of positive or negative linear correlation between the respective biomarkers.

## Notes

### Clinical Trial

NCT04478539

### Author Declarations

The local ethical committee of the Zan Mitrev Clinic reviewed and approved the clinical practice, treatment procedures described, and the results reported in this manuscript and approved the submission, #EBPZ.357. Trial registration: ClinicalTrials.gov, NCT04478539. Registered 14th of July 2020 - Retrospectively registered, https://clinicaltrials.gov/ct2/show/NCT04478539 Written informed (or temporary verbal) consent was obtained from all patients for publication of this manuscript and any accompanying images; the use of all health and medical information for scientific research and manuscript preparation was approved. A copy of the written consent is available for review by the Editor-in-Chief of this journal. For those patients that were not traceable after discharge, the need for informed consent was waived by the ethical committee.

## References

1. Huang C, Wang Y, Li X, Ren L, Zhao J, Hu Y, et al. Clinical features of patients infected with 2019 novel coronavirus in Wuhan, China. Lancet. 2020 Feb 15;395(10223):497–506.

2. Cucinotta D, Vanelli M. WHO Declares COVID-19 a Pandemic. Acta bio-medica : Atenei Parmensis. 2020 Mar 19;91(1):157–60.

3. Fara A, Mitrev Z, Rosalia RA, Assas BM. Cytokine storm and COVID-19: a chronicle of pro-inflammatory cytokines. Open Biology. 2020;10(9):200160.

4. Lucas C, Wong P, Klein J, Castro TBR, Silva J, Sundaram M, et al. Longitudinal analyses reveal immunological misfiring in severe COVID-19. Nature. 2020 2020/08/01;584(7821):463–69.

5. Shi Y, Wang Y, Shao C, Huang J, Gan J, Huang X, et al. COVID-19 infection: the perspectives on immune responses. Cell Death & Differentiation. 2020 2020/03/23.

6. Cleverley J, Piper J, Jones MM. The role of chest radiography in confirming covid-19 pneumonia. BMJ. 2020;370:m2426.

7. Yang L, Liu S, Liu J, Zhang Z, Wan X, Huang B, et al. COVID-19: immunopathogenesis and Immunotherapeutics. Signal Transduction and Targeted Therapy. 2020 2020/07/25;5(1):128.

8. Zhao Y, Nie H-X, Hu K, Wu X-J, Zhang Y-T, Wang M-M, et al. Abnormal immunity of non-survivors with COVID-19: predictors for mortality. Infectious Diseases of Poverty. 2020 2020/08/03;9(1):108.

9. Zhou F, Yu T, Du R, Fan G, Liu Y, Liu Z, et al. Clinical course and risk factors for mortality of adult inpatients with COVID-19 in Wuhan, China: a retrospective cohort study. Lancet. 2020 Mar 11.

10. Lindner D, Fitzek A, Bräuninger H, Aleshcheva G, Edler C, Meissner K, et al. Association of Cardiac Infection With SARS-CoV-2 in Confirmed COVID-19 Autopsy Cases. JAMA Cardiology. 2020.

11. Puntmann VO, Carerj ML, Wieters I, Fahim M, Arendt C, Hoffmann J, et al. Outcomes of Cardiovascular Magnetic Resonance Imaging in Patients Recently Recovered From Coronavirus Disease 2019 (COVID-19). JAMA Cardiology. 2020.

12. Ruan Q, Yang K, Wang W, Jiang L, Song J. Clinical predictors of mortality due to COVID-19 based on an analysis of data of 150 patients from Wuhan, China. Intensive Care Medicine. 2020 2020/03/03.

13. Connors JM, Levy JH. COVID-19 and its implications for thrombosis and anticoagulation. Blood. 2020;135(23):2033–40.

14. The Lancet H. COVID-19 coagulopathy: an evolving story. The Lancet Haematology. 2020;7(6):e425.

15. Petar Ugurov, Dijana Popevski, Tanja Gramosli, Dashurie Neziri, Dragica Vuckova, Marko Gjorgon, et al. Early Initiation of Extracorporeal Blood Purification Using the AN69ST (oXiris®) Hemofilter as a Treatment Modality for COVID-19 Patients: a Single-Centre Case Series. Braz J Cardiovasc Surg. 2020.

16. Spagnolo P, Balestro E, Aliberti S, Cocconcelli E, Biondini D, Casa GD, et al. Pulmonary fibrosis secondary to COVID-19: a call to arms? The Lancet Respiratory Medicine. 2020;8(8):750–52.

17. Wunsch H. Mechanical Ventilation in COVID-19: Interpreting the Current Epidemiology. Am J Respir Crit Care Med. 2020;202(1):1–4.

18. Peng PWH, Ho P-L, Hota SS. Outbreak of a new coronavirus: what anaesthetists should know. British Journal of Anaesthesia.

19. Cennimo DJ. Coronavirus Disease 2019 (COVID-19) Guidelines: CDC Interim Guidance on Coronavirus Disease 2019 (COVID-19). 2020.

20. Commission TGOoNH. Diagnosis and Treatment Protocol for Novel Coronavirus Pneumonia. 2020.

21. Poston JT, Patel BK, Davis AM. Management of Critically Ill Adults With COVID-19. JAMA. 2020.

22. Coronavirus drugs trials must get bigger and more collaborative. Nature. 2020 2020-05-13;581(7807):120–20.

23. Health NIo. Guidelines Introduction | Coronavirus Disease COVID-19. @NIHCOVIDTxGuide; 2020.

24. Ngo BT, Marik P, Kory P, Shapiro L, Thomadsen R, Iglesias J, et al. A SYSTEMATIC ANALYSIS OF THE TIME COURSE TO DEVELOP TREATMENTS FOR COVID-19. medRxiv. 2020:2020.05.27.20115238.

25. Riva L, Yuan S, Yin X, Martin-Sancho L, Matsunaga N, Pache L, et al. Discovery of SARS-CoV-2 antiviral drugs through large-scale compound repurposing. Nature. 2020 2020/07/24.

26. Rieder M, Zahn T, Benk C, Lother A, Bode C, Staudacher D, et al. Cytokine adsorption in a patient with severe coronavirus disease 2019 related acute respiratory distress syndrome requiring extracorporeal membrane oxygenation therapy: A case report. Artificial Organs.n/a(n/a).

27. Zhang H, Zhu G, Yan L, Lu Y, Fang Q, Shao F. The absorbing filter Oxiris in severe coronavirus disease 2019 patients: A case series. Artificial Organs.n/a(n/a).

28. Al Shareef K, Bakouri M. Cytokine Blood Filtration Responses in COVID-19. Blood Purification. 2020.

29. Damiani M, Gandini L, Landi F, Fabretti F, Gritti G, Riva I. Extracorporeal Cytokine Hemadsorption in Severe COVID-19 Respiratory Failure. medRxiv. 2020:2020.06.28.20133561.

30. Rieder M, Wengenmayer T, Staudacher D, Duerschmied D, Supady A. Cytokine adsorption in patients with severe COVID-19 pneumonia requiring extracorporeal membrane oxygenation. Critical Care. 2020 2020/07/14;24(1):435.

31. Villa G, Zaragoza JJ, Sharma A, Neri M, De Gaudio AR, Ronco C. Cytokine removal with high cut-off membrane: review of literature. Blood Purif. 2014;38(3-4):167–73.

32. Malard B, Lambert C, Kellum JA. In vitro comparison of the adsorption of inflammatory mediators by blood purification devices. Intensive care medicine experimental. 2018 May 4;6(1):12.

33. Chen G, Zhou Y, Ma J, Xia P, Qin Y, Li X. Is there a role for blood purification therapies targeting cytokine storm syndrome in critically severe COVID-19 patients? Renal failure. 2020 Nov;42(1):483–88.

34. Broman ME, Hansson F, Vincent JL, Bodelsson M. Endotoxin and cytokine reducing properties of the oXiris membrane in patients with septic shock: A randomized crossover double-blind study. PloS one. 2019;14(8):e0220444.

35. Kogelmann K, Jarczak D, Scheller M, Drüner M. Hemoadsorption by CytoSorb in septic patients: a case series. Critical care (London, England). 2017 Mar 27;21(1):74.

36. Monard C, Rimmelé T, Ronco C. Extracorporeal Blood Purification Therapies for Sepsis. Blood Purif. 2019;47 Suppl 3:1–14.

37. Gómez-Pastora J, Weigand M, Kim J, Wu X, Strayer J, Palmer AF, et al. Hyperferritinemia in critically ill COVID-19 patients - Is ferritin the product of inflammation or a pathogenic mediator? Clin Chim Acta. 2020;509:249–51.

38. Velavan TP, Meyer CG. Mild versus severe COVID-19: Laboratory markers. International Journal of Infectious Diseases. 2020 2020/06/01/;95:304–07.

39. Pourcyrous M, Korones SB, Crouse D, Bada HS. Interleukin-6 (IL-6) and C-reactive protein (CRP) Responses to Immunization in Premature Babies † 1007. Pediatric Research. 1997 1997/04/01;41(4):170–70.

40. McArdle PA, McMillan DC, Sattar N, Wallace AM, Underwood MA. The relationship between interleukin-6 and C-reactive protein in patients with benign and malignant prostate disease. British Journal of Cancer. 2004 2004/11/01;91(10):1755–57.

41. Panichi V, Manca-Rizza G, Paoletti S, Taccola D, Consani C, Filippi C, et al. Effects on inflammatory and nutritional markers of haemodiafiltration with online regeneration of ultrafiltrate (HFR) vs online haemodiafiltration: a cross-over randomized multicentre trial. Nephrology, dialysis, transplantation : official publication of the European Dialysis and Transplant Association - European Renal Association. 2006 Mar;21(3):756–62.

42. Wigmore SJ, Fearon KC, Maingay JP, Lai PB, Ross JA. Interleukin-8 can mediate acute-phase protein production by isolated human hepatocytes. The American journal of physiology. 1997 Oct;273(4):E720–6.

43. Gabay C, Kushner I. Acute-Phase Proteins and Other Systemic Responses to Inflammation. New England Journal of Medicine. 1999;340(6):448–54.

44. Sproston NR, Ashworth JJ. Role of C-Reactive Protein at Sites of Inflammation and Infection. Frontiers in immunology. 2018;9:754.

45. Weinhold B, Bader A, Poli V, Rüther U. Interleukin-6 is necessary, but not sufficient, for induction of the humanC-reactive protein gene in vivo. The Biochemical journal. 1997 Aug 1;325 (Pt 3)(Pt 3):617–21.

46. Del Valle DM, Kim-Schulze S, Huang H-H, Beckmann ND, Nirenberg S, Wang B, et al. An inflammatory cytokine signature predicts COVID-19 severity and survival. Nature Medicine. 2020 2020/08/24.

47. Kox M, Waalders NJB, Kooistra EJ, Gerretsen J, Pickkers P. Cytokine Levels in Critically Ill Patients With COVID-19 and Other Conditions. JAMA. 2020.

48. Sinha P, Matthay MA, Calfee CS. Is a “Cytokine Storm” Relevant to COVID-19? JAMA Internal Medicine. 2020;180(9):1152–54.

49. Ragab D, Salah Eldin H, Taeimah M, Khattab R, Salem R. The COVID-19 Cytokine Storm; What We Know So Far. Frontiers in immunology. 2020 2020-June-16;11(1446).

50. Chau AS, Weber AG, Maria NI, Narain S, Liu A, Hajizadeh N, et al. The Longitudinal Immune Response to Coronavirus Disease 2019: Chasing the Cytokine Storm. Arthritis & Rheumatology.n/a(n/a).

51. Chen LYC, Hoiland RL, Stukas S, Wellington CL, Sekhon MS. Confronting the controversy: Interleukin-6 and the COVID-19 cytokine storm syndrome. European Respiratory Journal. 2020:2003006.

52. Wang J, Li Q, Yin Y, Zhang Y, Cao Y, Lin X, et al. Excessive Neutrophils and Neutrophil Extracellular Traps in COVID-19. Frontiers in immunology. 2020 2020-August-18;11(2063).

53. Didangelos A. COVID-19 Hyperinflammation: What about Neutrophils? mSphere. 2020;5(3):e00367–20.

54. Drent M, Cobben NA, Henderson RF, Wouters EF, van Dieijen-Visser M. Usefulness of lactate dehydrogenase and its isoenzymes as indicators of lung damage or inflammation. The European respiratory journal. 1996 Aug;9(8):1736–42.

55. Kishaba T, Tamaki H, Shimaoka Y, Fukuyama H, Yamashiro S. Staging of acute exacerbation in patients with idiopathic pulmonary fibrosis. Lung. 2014Feb;192(1):141–9.

56. Yan L, Zhang H-T, Goncalves J, Xiao Y, Wang M, Guo Y, et al. An interpretable mortality prediction model for COVID-19 patients. Nature Machine Intelligence. 2020 2020/05/01;2(5):283–88.

57. Langford BJ, So M, Raybardhan S, Leung V, Westwood D, MacFadden DR, et al. Bacterial co-infection and secondary infection in patients with COVID-19: a living rapid review and meta-analysis. Clinical Microbiology and Infection.

58. Sieswerda E, De Boer MGJ, Bonten MMJ, Boersma WG, Jonkers RE, Aleva RM, et al. Recommendations for antibacterial therapy in adults with COVID-19 – An evidence based guideline. Clinical Microbiology and Infection.

59. Contou D, Claudinon A, Pajot O, Micaëlo M, Longuet Flandre P, Dubert M, et al. Bacterial and viral co-infections in patients with severe SARS-CoV-2 pneumonia admitted to a French ICU. Annals of Intensive Care. 2020 2020/09/07;10(1):119.

60. van den Bosch CM, Hulscher ME, Natsch S, Gyssens IC, Prins JM, Geerlings SE. Development of quality indicators for antimicrobial treatment in adults with sepsis. BMC infectious diseases. 2014 Jun 20;14:345.

61. WHO. Clinical management of severe acute respiratory infections (SARI) when COVID-19 disease is suspected. World Health Organization Geneva; 2020.

62. Adler H, Ball R, Fisher M, Mortimer K, Vardhan MS. Low rate of bacterial co-infection in patients with COVID-19. The Lancet Microbe. 2020;1(2):e62.

63. Versporten A, Bolokhovets G, Ghazaryan L, Abilova V, Pyshnik G, Spasojevic T, et al. Antibiotic use in eastern Europe: a cross-national database study in coordination with the WHO Regional Office for Europe. The Lancet Infectious diseases. 2014 May;14(5):381–7.

64. Horvat O, Mijatović V, Milijasević B, Tomas A, Kusturica MP, Tomić Z, et al. Are There Striking Differences in Outpatient Use of Antibiotics Between South Backa District, Serbia, and Some Scandinavian Countries? Frontiers in public health. 2018;6:91.

65. Jakupi A, Raka D, Kaae S, Sporrong SK. Culture of antibiotic use in Kosovo - an interview study with patients and health professionals. Pharmacy practice. 2019 Jul-Sep;17(3):1540.

66. Susan van den Hof SWJMKKIWDLFWSNMGOKAT. Central Asian and European Surveillance of Antimicrobial Resistance. Annual report 2019. 2019 2019-11-18.

67. Chen Y-T, Shao S-C, Hsu C-K, Wu IW, Hung M-J, Chen Y-C. Incidence of acute kidney injury in COVID-19 infection: a systematic review and meta-analysis. Critical care (London, England). 2020;24(1):346–46.

68. Quah P, Li A, Phua J. Mortality rates of patients with COVID-19 in the intensive care unit: a systematic review of the emerging literature. Critical care (London, England). 2020 Jun 4;24(1):285.

