## Supplementary figures and images for "Extracorporeal Blood Purification in moderate and severe COVID-19 patients: a prospective cohort study"

### supplemental figure 1

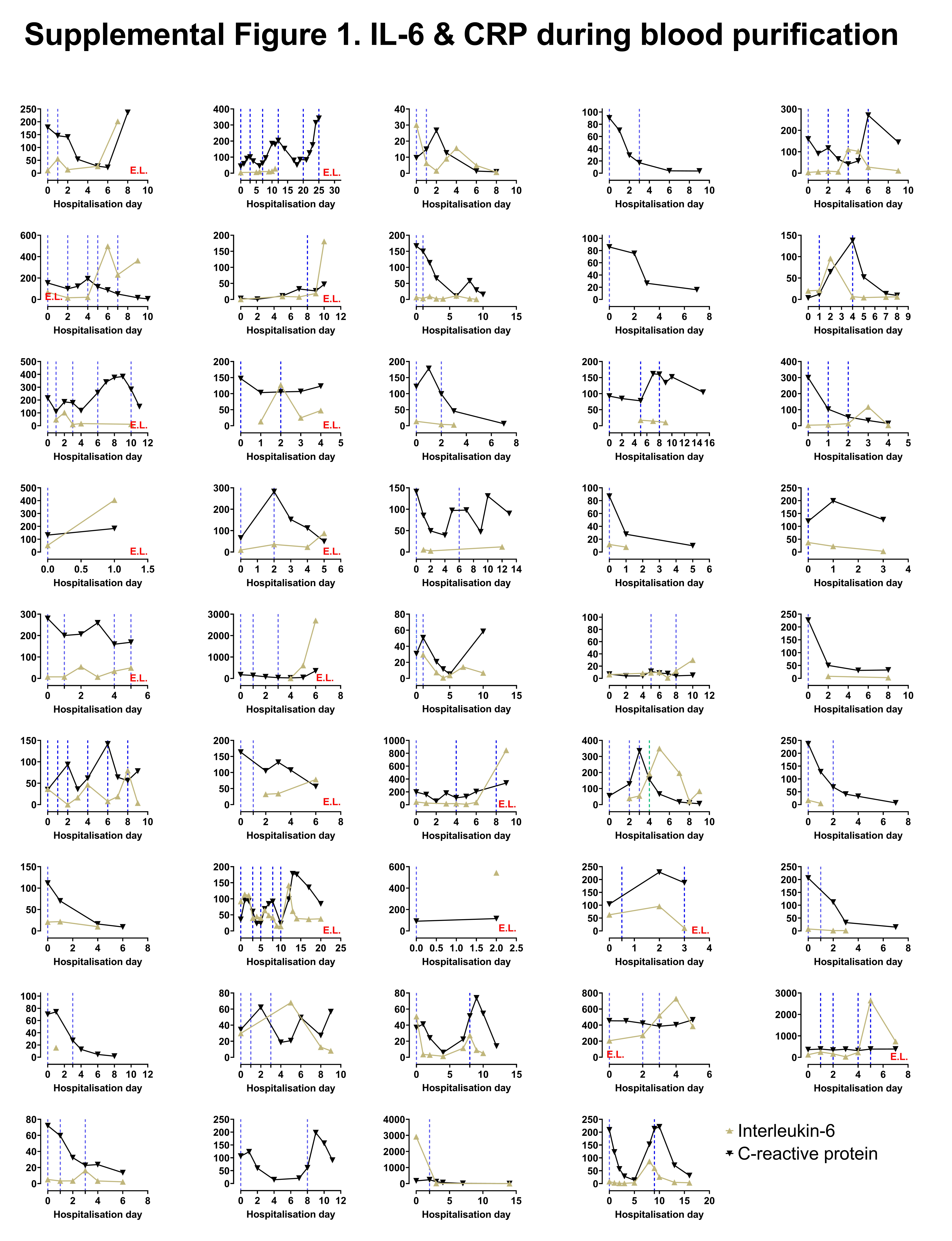

### supplemental figure 2

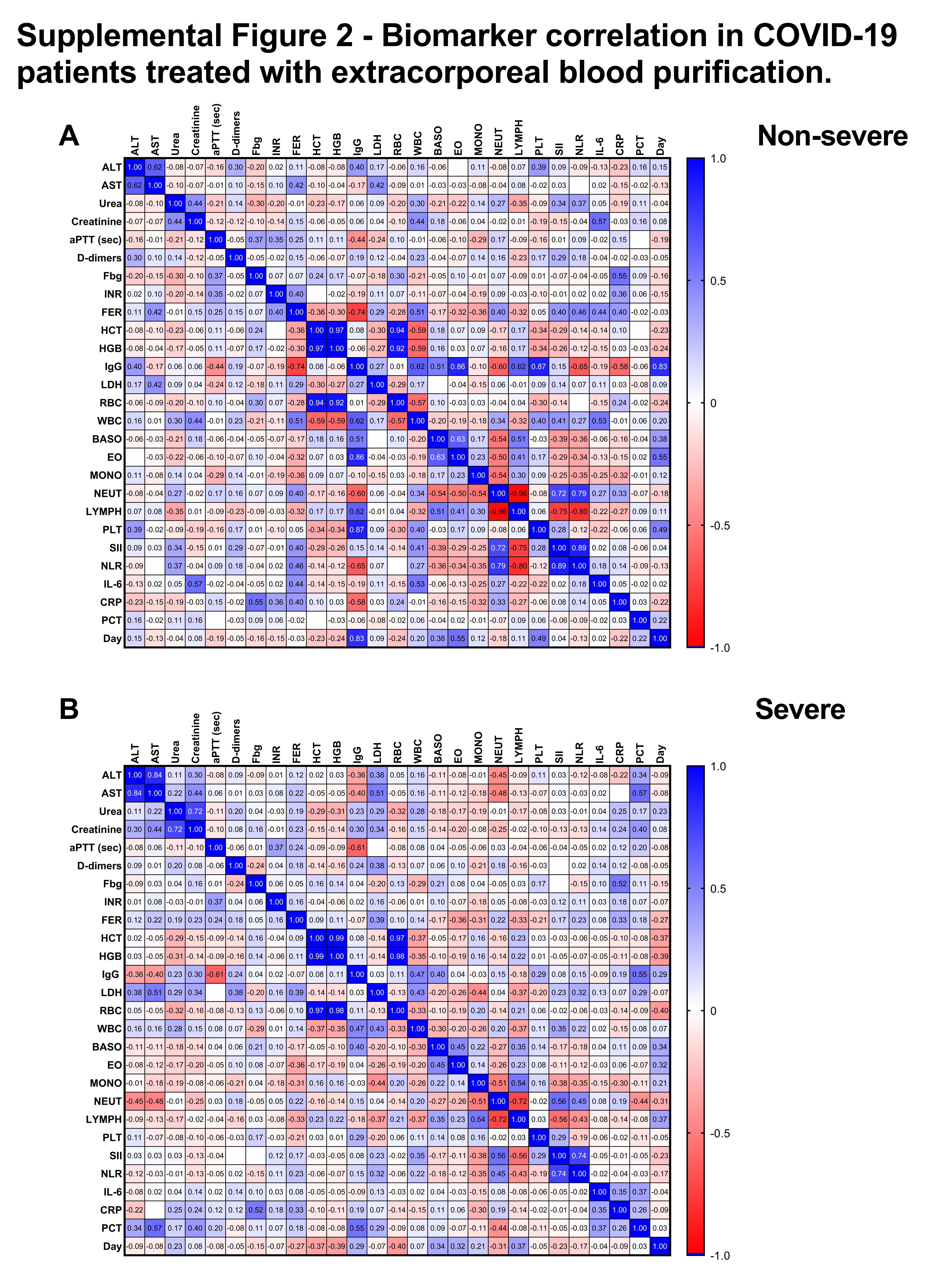
